# Risk Analysis and Prediction for COVID19 Demographics in Low Resource Settings using a Python Desktop App and Excel Models

**DOI:** 10.1101/2020.04.13.20063453

**Authors:** Ali Kyagulanyi, Joel Tibabwetiza Muhanguzi, Dembe Oscar

**Affiliations:** Makere University Kampala; Makerere University; Makerere Univesity

## Abstract

While the novel covid19 disease caused by sar-cov-2 corona virus has proved a serious threat to mankind it being a pandemic, the rate at which technology in low resource income countries like Uganda has been used to predict the spread and impact of the disease in their economies has not been strongly employed. This paper presents a an excel model and desktop application software developed using open source python programming tools for carrying out risk analysis and prediction of demographics for covid19 disease. Prediction results for both models clearly stated using epidemiological curve, these results can vary based on the force of infection which varies based on government measures and actions. With a certain degree of certainty of the potential impact of the disease on low resource countries, it will foster proper planning and strategical methods to properly manage the pandemic.

## 1.0 INTRODUCTION

The early January outbreak of the novel covid19 disease in Wuhan city and Hubei provinces in China caught the world unaware and got scientists, governments and all the people around the globe working at extremely high pressure and speed. By 28^th^ march 2020 the virus had claimed more than 28238 lives, 614063 infections in most of the countries around the world according to the world health organization. The very fast spread of the disease has led to a declaration of a pandemic by the world health organization. The United States of America (USA) currently has 104,256 infections with 1074 deaths which is the highest number currently worldwide. Italy follows in line with 9314 deaths and 86498 cases. China currently in 3^rd^ place, claimed the biggest share of the initial cases with more than 81394 infections and 3295 deaths. World Health Organization (WHO) confirmed that covid19 disease is deadlier than seasonal flue with a global mortality rate at 3.4% but doesn’t transmit as easily (times 2020).

Although the virus originated in China, WHO has declared Europe as the epicenter of the disease after China. Africa is the only content to have fewer cases and people with confirmed cases. In just ten days, the virus has spread to more than 16 countries in Africa, with most cases confirmed to be imported. Egypt has the highest number of cases 109 and 1 death.

According to the Ministry of Health in Uganda, there are currently 30 confirmed cases of covid19. However, tensions remain high in the East African Community as neighboring countries Kenya, DRC and Rwanda have reported more cases. Governments in developed countries have invested a great amount of funds and put in place strict measures to safe guard their citizens, the biggest scare lies in low resource countries like Uganda, and WHO believes that African countries are not ready to battle the disease because of the undependable health care system. Planning and preparation of an inevitable outbreak has been tricky in Uganda as statistical prediction has not been fully exploited to determine the likely impact of the disease to the Ugandan population. This is the case for most of the developing countries’ health care systems across the globe.

Governments around the world have found it hard to estimate the potential burden of the pandemic, hence countries like Uganda have not yet fully established a contingency plan based on statistical and mathematical predictions. This has led to low efforts in expanding and sustaining investment to build preparedness and health capacity in Uganda. Using Italy and South Korea as an example, South Korea confirmed its first case on 20^th^ January (Wikipedia 2020) and by 22^nd^ march South Korea has 2909 cases with only 104 deaths. On the other hand, Italy confirmed its first cases on 31^st^ January 2020 and has 86498 cases with 9314 deaths by 28^th^ march 2020 (University 2020). This was due to under planning for the pandemic which has registered the most severe effects compared to a country like South Korea which planned resources based on mathematical and computational predictions. South Korea employed mathematical predictions like kinetic model which was used to simplify the SIR model to plan (GeneOnline 2020), for the number of patients and make key decisions on quarantines’ and city lock down, For South Korea this seems have paid off as they have managed to steadily suppress the pandemic.

This paper presents a risk analysis and statistical prediction Desktop application and excel model of covid19 disease to the populations in low resource settings countries like Uganda. It predicts the trends in case of an outbreak for a period of at most 60 days from onset. The algorithm in the application employs a mathematical model of an Ordinary Differential Equation representing the SIR model (susceptible, infected and recovered (or removed) which has been used by epidemiologists in the past to predict the number of individuals likely to be infected, recover or die in the event of any epidemic. The models give results for expected number of infections in a single susceptible district like Kampala. Using infection force from different countries we sampled two rates of infection 0.29 and 0.594 for moderate and worst-case scenarios and generated epidemiological curves showing the trend the disease is expected to follow.

Similar models have been designed to predict the fate of the disease in countries like united states of America, United Kingdom and South Korea and the have been found to have a high predictability. The application accepts inputs of force of infection, recovery, number of people in an area, initial number of people infected and recovered from the population. Using the python programming language, calculus and condition probability we have designed the algorithm that generates epidemiological curves and parameter (SIR) tables as an output, the efficiency of the application has been tested with a similar model implemented in an excel worksheet, predictions and graphs have been made from excel.

## 2.0 OBJECTIVES

- To design and implement a desktop application (python model) and excel model that predicts for covid19 disease in Uganda that predicts the behavior of the pandemic for the first 60 days on the day of onset
- To predict and give statistical values and graphs of people who are likely to be infected, recover and die from the disease.
- To design a software that automatically performs a risk analysis and progress as the fight against the disease intensifies as the number of cases increase in the country and generates automatic epidemiological curves.

## 3.0 LITERATURE REVIEW

SIR model, also commonly known as compartmental modelling, is a technique used to simplify the mathematical analysis of an infectious disease. It has been used to predict the future trends of diseases that can be transmitted from human to human, like measles, rubella and mumps (REF). The desired population is divided into compartments susceptible infected and recovered. Each individual in the entire population(N) is assigned to any of the three compartments. Susceptible individuals have no immunity to the disease and they can therefore be infected through exposure to the virus or after contact with an infectious person, susceptible individuals can move to infectious after the infection. Infectious people have the disease and can spread it to others, once infectious people recovery they move to recovered. Since people can move between the three compartments, the number of people in each compartment changes over time.

The rate of change of individuals in the three regions with time follows an SIR model. Mathematically the SIR model will capture population changes in each compartment with a system of ordinary differential equations (ODEs) to model the progression of the disease. To get a clearer prediction we have to temporarily ignore the natural birth and death rate in the population, the ODE follows the equation below.

### 3.1 PARAMETERS

#### Ro (pronounced R-naught)

Ro is essentially a metric of how contagious a disease is. In simpler terms, Ro is the average number of people in a susceptible population that a single infected person will spread the disease to over the course of their infection. Ro can capture three basic scenarios:

If Ro is less than one, on average, an infected person infects less than one person and disease is expected to stop spreading.

If Ro is equal to one an infected person infects an average of one person, the disease spread is stable, or endemic, and the number of infections is not expected to increase or decrease.

If Ro is greater than one on average, an infected person infects more than one person, the disease is expected to increasingly spread in the absence of intervention.

Early estimates estimate the value of covide19 to be between 2 and 6 based on rate of contact and population of the area.

#### 3.1.1 ORDINARY DIFFERENTIAL EQUATION

SIR model assumed that the rate of change of susceptibility, recovery and infection with time follows an ordinary differential equation with three variables whose sum is equal to zero. On solving the equation, the variables should equal to a constant N which is the Number of people in a population.

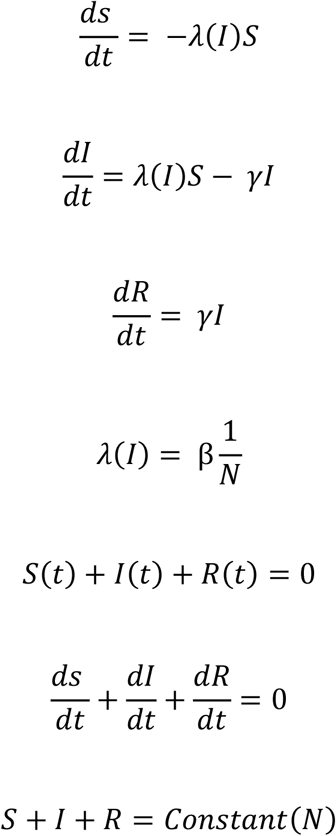

Where:

λ is the rate at which susceptible individuals become infectious—called the force of infection.

γ is the recovery rate, the rate at which people recover from infection. This is the time someone spends while infecting other people in the population. For covid19 it is estimated to be between 14 to 25 days, for worst case scenario (Ugandan case) gamma is taken to be the extreme 25 hence 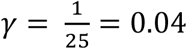

β, the transmission rate − the product of the rate of contact and the probability of transmission given contact. This also known as the force of infection. Transmission rate (rate of infection) depends on the action taken by the government to safe guard the population against corona virus and is determined using either the reproductive number or information obtained from previous analysis. For this paper we have used three estimates of rate of infection. Different online publications have estimated beta to be 0.1 (Scott P. Layne 11 Mar 2020:), 0.28 and 0.594. Where 0.1 is usually obtained for a lower reproducibility number and it is used when the government has taken strict measures to safe guard the population from the pandemic we have referred to this as maximum government action. 0.28 is obtained when the population is taking moderate measures to fight against the disease, this is the infection rate for countries in. The following values for infection rate are shown in the table below and we have used them as inputs to obtain result and three case scenarios of the predictions.

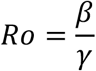

| Constants | Moderate | No action |
| --- | --- | --- |
| Beta (B) | 0.28 | 0.594 |
| Gamma ($\gamma$) | 0.04 | 0.04 |

The input parameters shall be constant for all the different areas in Uganda, as shown in the table below for ca chosen number of districts. For the total population who are likely to be infected, we have sampled three major cities in Uganda obtaining their populations for 2020 as projected by Uganda national bureau of statistics. The sampled Cities include Kampala, Wakiso, Kalangala districts. We the sampled Uganda as a whole, using the entire population as an input as shown in the table below (STATISTIC 2019).

| Region (District) | Population |
| --- | --- |
| Kampala | 1,650,800 |
| Wakiso | 2,080,00 |

### 3.2 METHOD ONE: DESKTOP APPLICATION

The desktop application was developed using open source software tools. It was designed in line with the agile software development lifecycle. This ensured a timely software modification during the development process.

Requirements analysis was done for the software to establish both functional and non-functional requirements. The figure below shows the structure of the prediction and risk analysis software.

**Figure 1.**
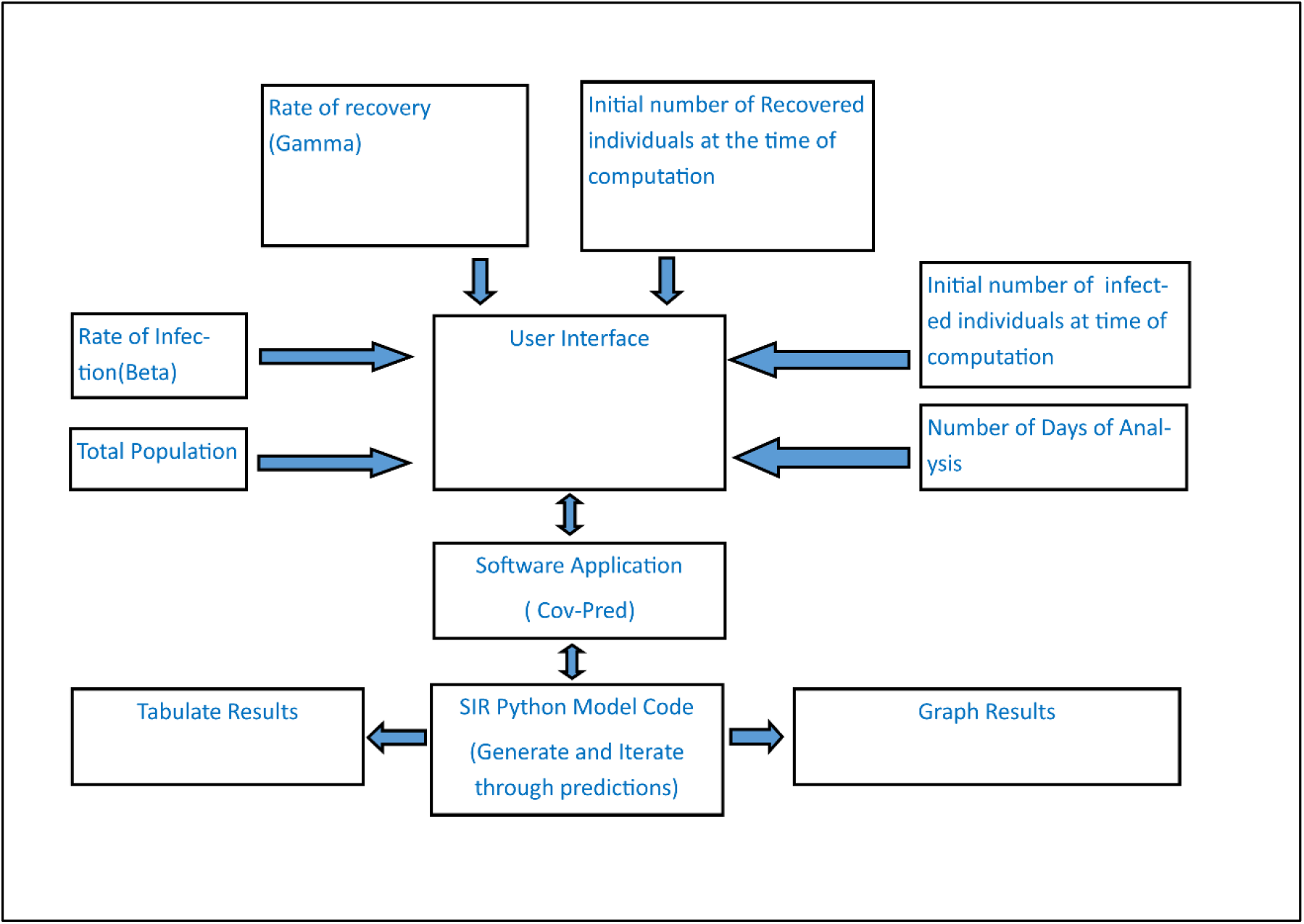
Software Block Diagram

The software uses python as the core programming language. This enables to take advantage of the vast collection of python’s open source libraries which are described below.

- matplotlib that is used to generate graphical representations of data predictions based on the SIR epidemic model.
- The Scipy library is used to calculate the predictions based on ordinary differential equations.
- The tkinter and kivy libraries are used to construct a user-friendly interface with the software through which inputs are subjected to the software which can then use to compute the predictions of the number of people based on demographics.

**Figure 2:**
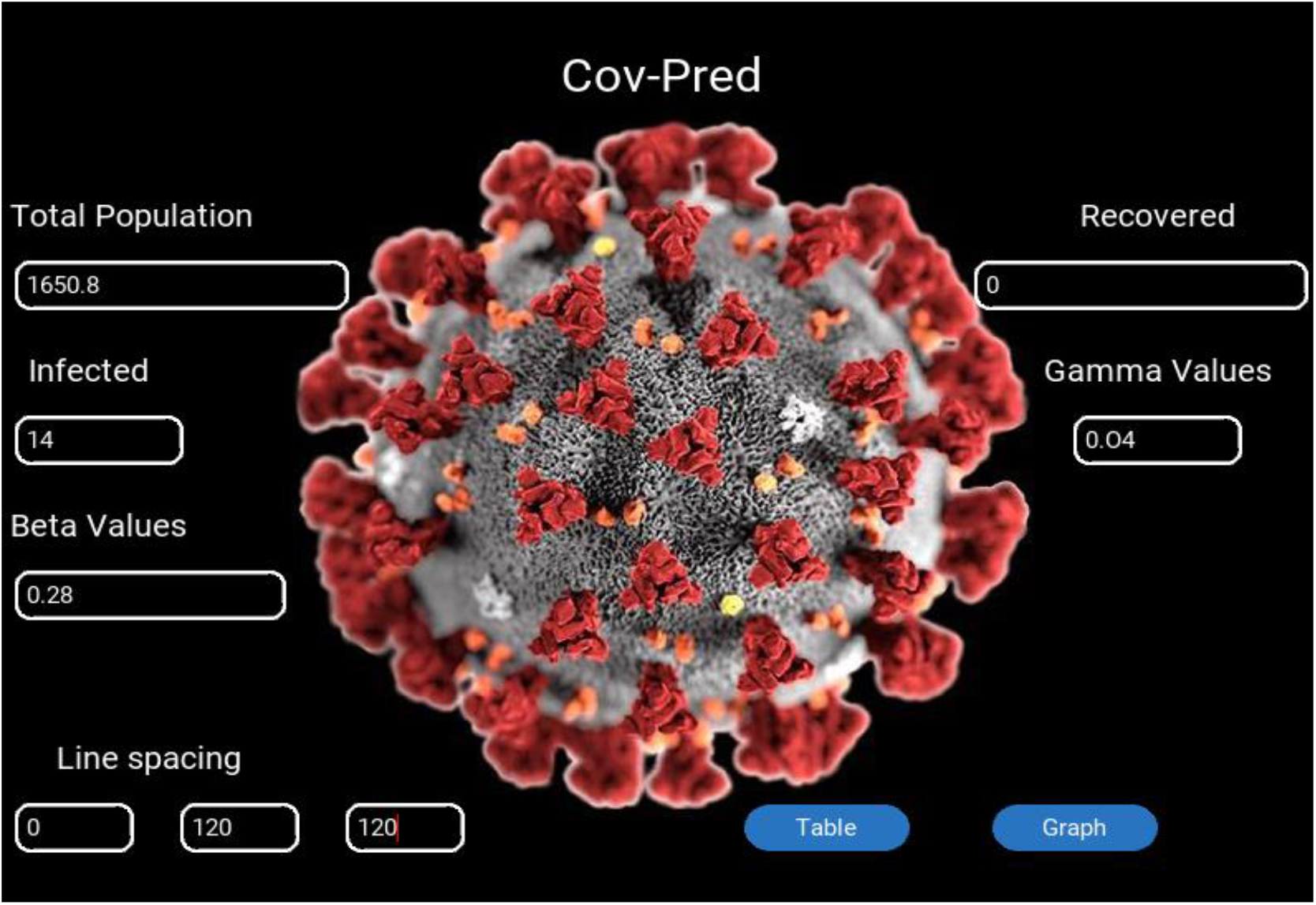
Lay out of the predictive application

The software utilizes 6 main parameters as shown in the figure above.

- total population of a given area i.e. district or town, the population demographics based on the age
- the number of infected people prior to computation,
- the number of recovered individuals prior to computation,
- the current rate of infection of the epidemic as observed from other existing affected regions,
- the rate of recovery
- and the number of days for which the prediction is to be done.

The underlying algorithm model also depends on 3 main assumptions:

- The population remains constant through the time of prediction
- The total population is divided into three main compartments namely; susceptible, infected and removed, and an individual should belong to only one of them at any given time. Removed implies that an individual has either recovered or died. An individual can only move from susceptible to infected, then removed according to the SIR model and not vice versa.
- The rate of recovery and the rate of infection is constant.

#### 3.2.3 RESULTS

The figure below shows the graphical representation generated by the software, of the three compartments comprising the population according to the SIR model. This is based on the population demographics of Wakiso District. The code can be accessed [here](https://github.com/covpredapp/pymlcode)

**Figure 4.**
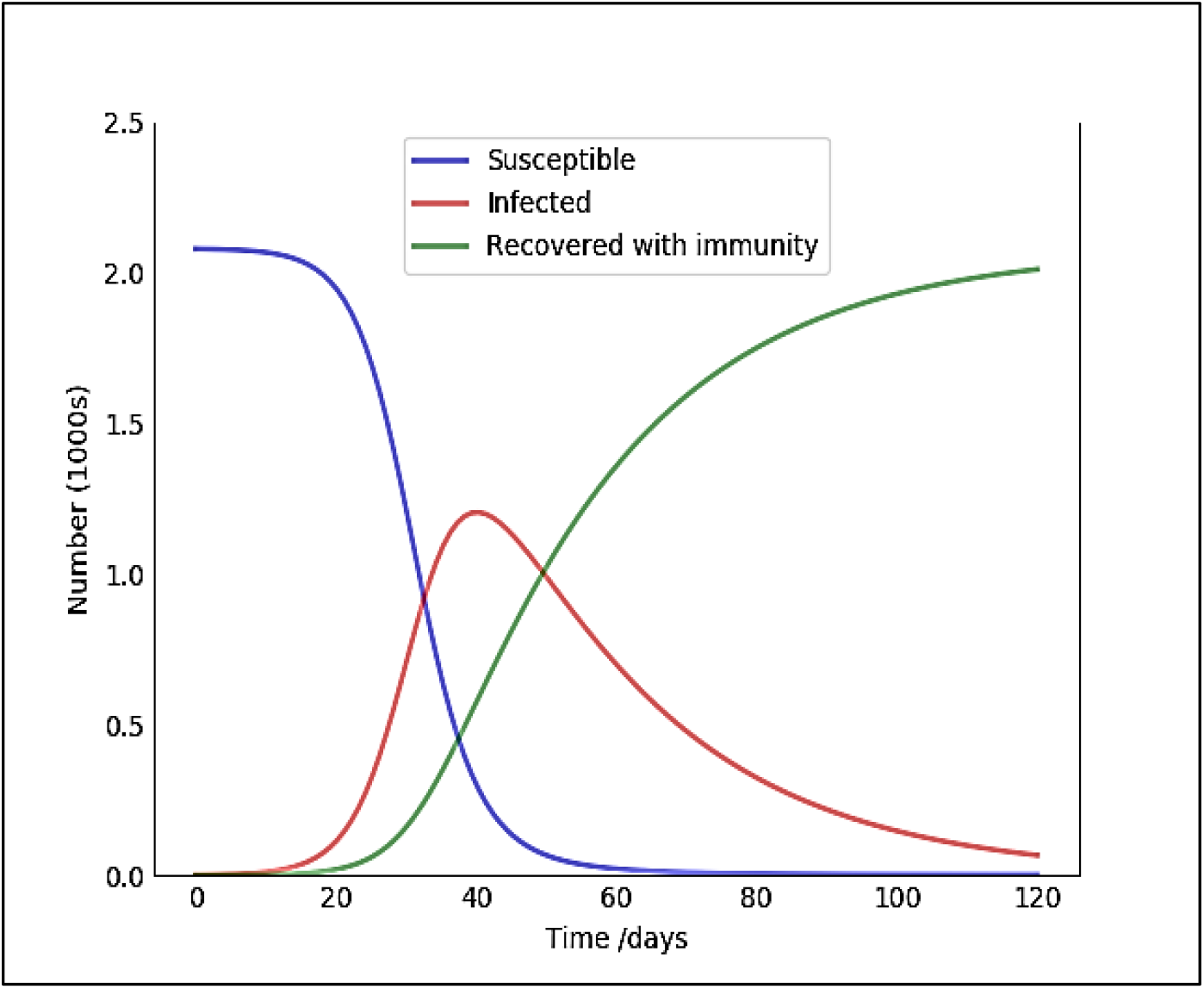
Wakiso District Graphical Predictions At B=0.28

The figure below illustrates the structure of the tabular prediction results generated by the desktop application per day from the first day of infection upto the specified future date which can also be exported in a csv file for further analysis.

**Figure 5:**
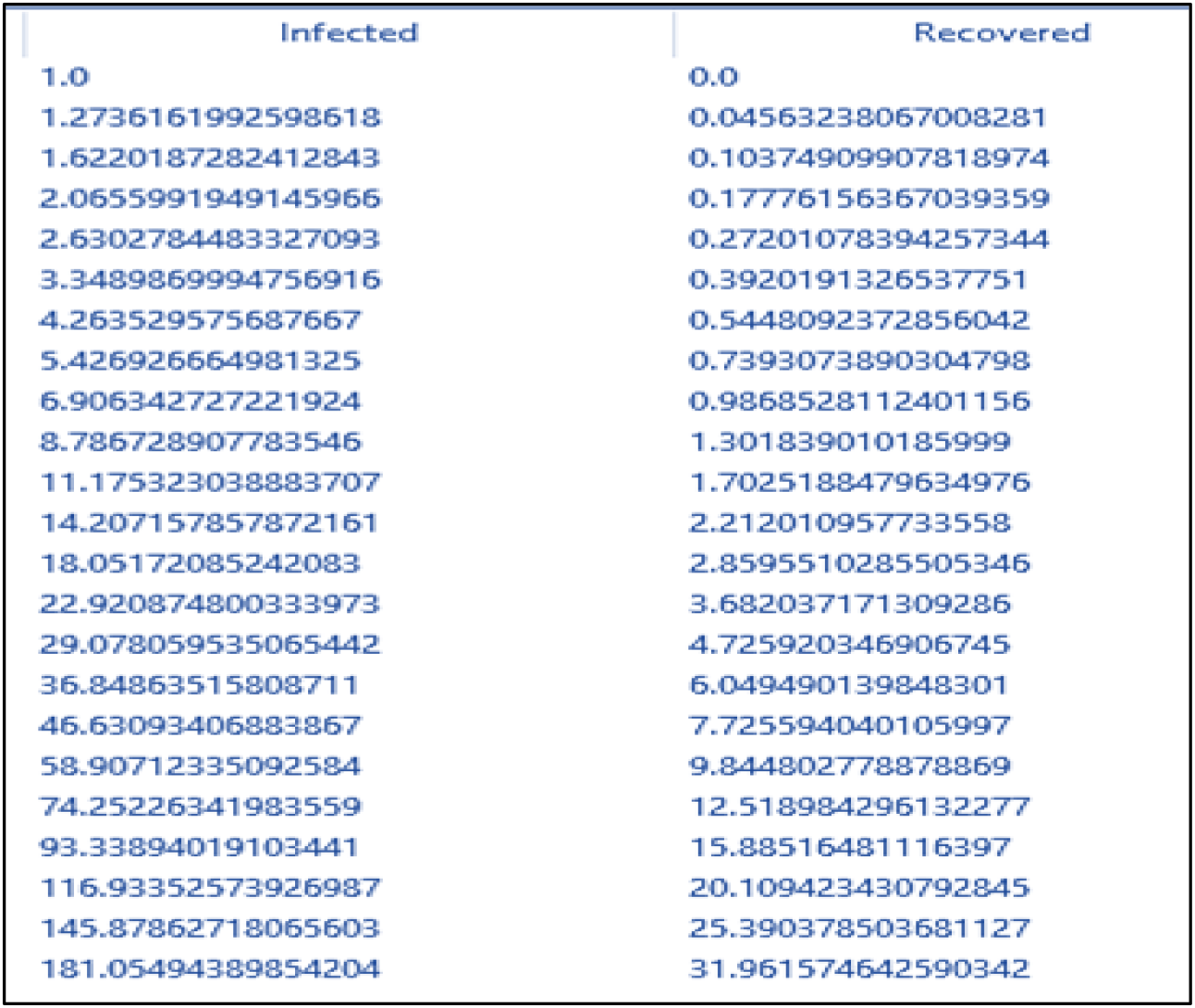
Table showing prediction results generated by app for people in wakiso district

### 3.3 METHOD TWO: EXCEL MODEL

In the excel model we have remodeled the SIR differential equation in an excel work sheet and compared. We modelled the same SIR ordinary differential equation input parameters for covid19 disease, this second model can be used as control and it will verify the results of the python algorithm. Both models use the same values of beta, gamma and population of a given area. We have also included the possible number of deaths in this model values of susceptible, infectious and recovered populations can be calculated using the mathematical equations below which are obtained after integration of the ordinary differential equations. They calculate the number of people in each condition today (n), based on the number yesterday (n-1) and the rates of change, ß and γ. The subscript n means the number in one-time interval, and n-1 means the number in the previous interval.

Equation one:

The number of susceptible people today (S) equals the number yesterday (S), minus the percentage of people who become infected today (which is yesterday’s number of susceptible people (S) divided by the original number (/S)), times their rate of infection (ß), times how many people were infected (I) yesterday.

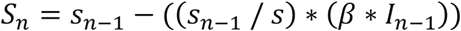

Equation two.

The number of infected people today (I) equals the number who were infected yesterday (I), plus the number of susceptible people who became infected today, minus the number of infected yesterdays who recovered.

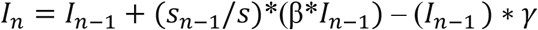

Equation three:

The number of recovered people today (R) equals the previous number who had recovered, plus the number who of infected people yesterday who recovered today.

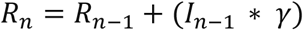

We have added the following three equations in an excel worksheet using excel formulas to model fit them in excel. These equations by hand for each day but the excel. We have calculated and tabulated results for the three excel equations in an excel spread sheet. On the left corner we have made a small table of variables, so that variables can easily be changed to see the effect on our model, see A5 to B12 in the Excel image below

#### 3.3.1 EQUATIONS EXCEL CELL FORMATS

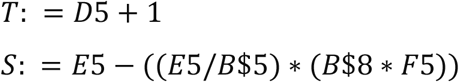

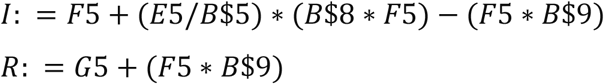

these equations not used for line D5 to H5 because Line D5 to H5 give the initial conditions

**Figure 6:**
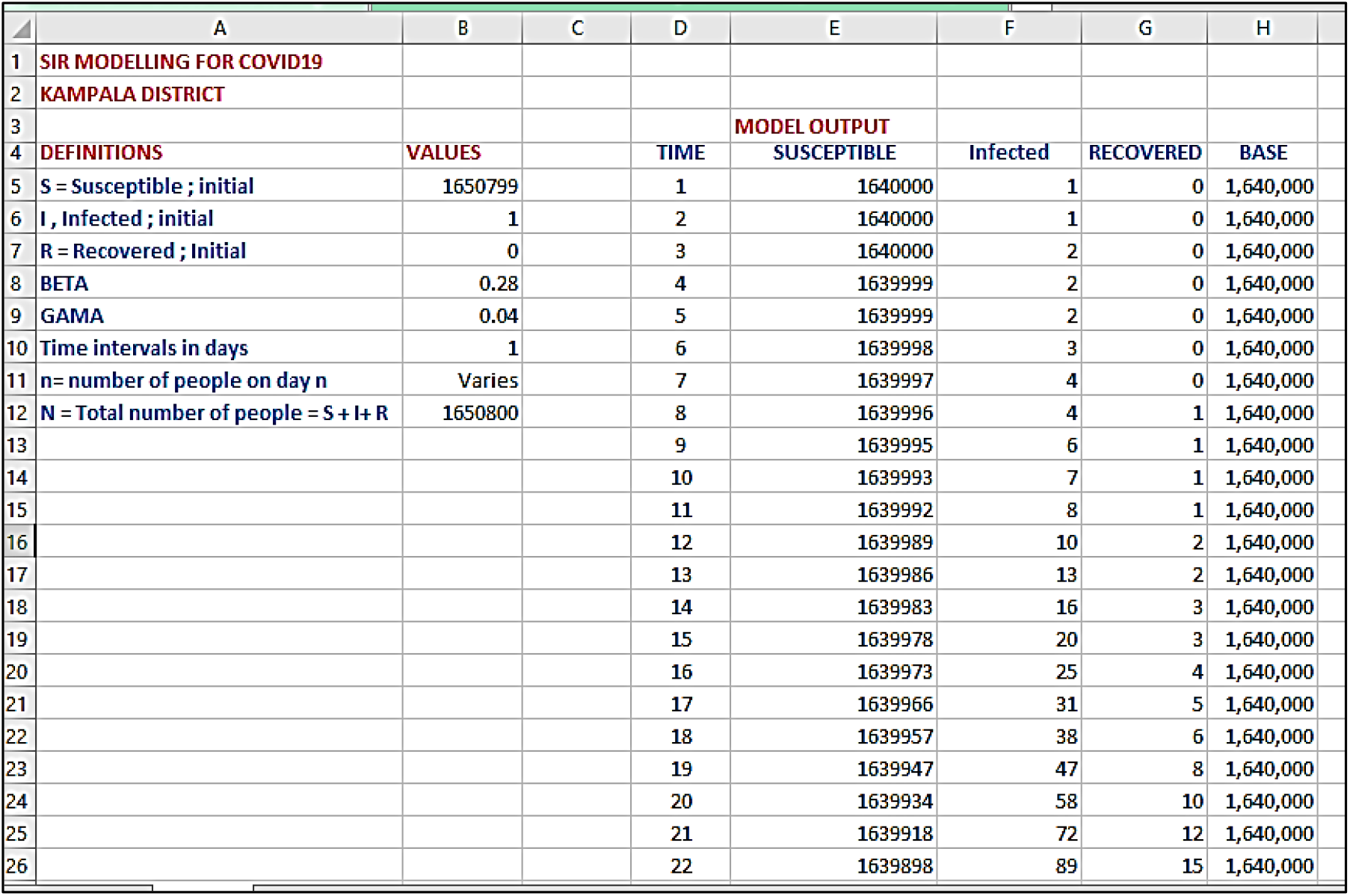
Shows the excel work sheet SIR model with inputs (blue) on the left and outputs on the right

#### 3.3.2 EXCEL RESULTS FOR KAMPALA

The two graphs have been generated from excel work sheets for the region of Kampala where the values of B have been varied between 0.28 and 0.594, which are the possible rates of infections picked from other countries. The test is done using Kampala region as a reference.

**Figure 7:**
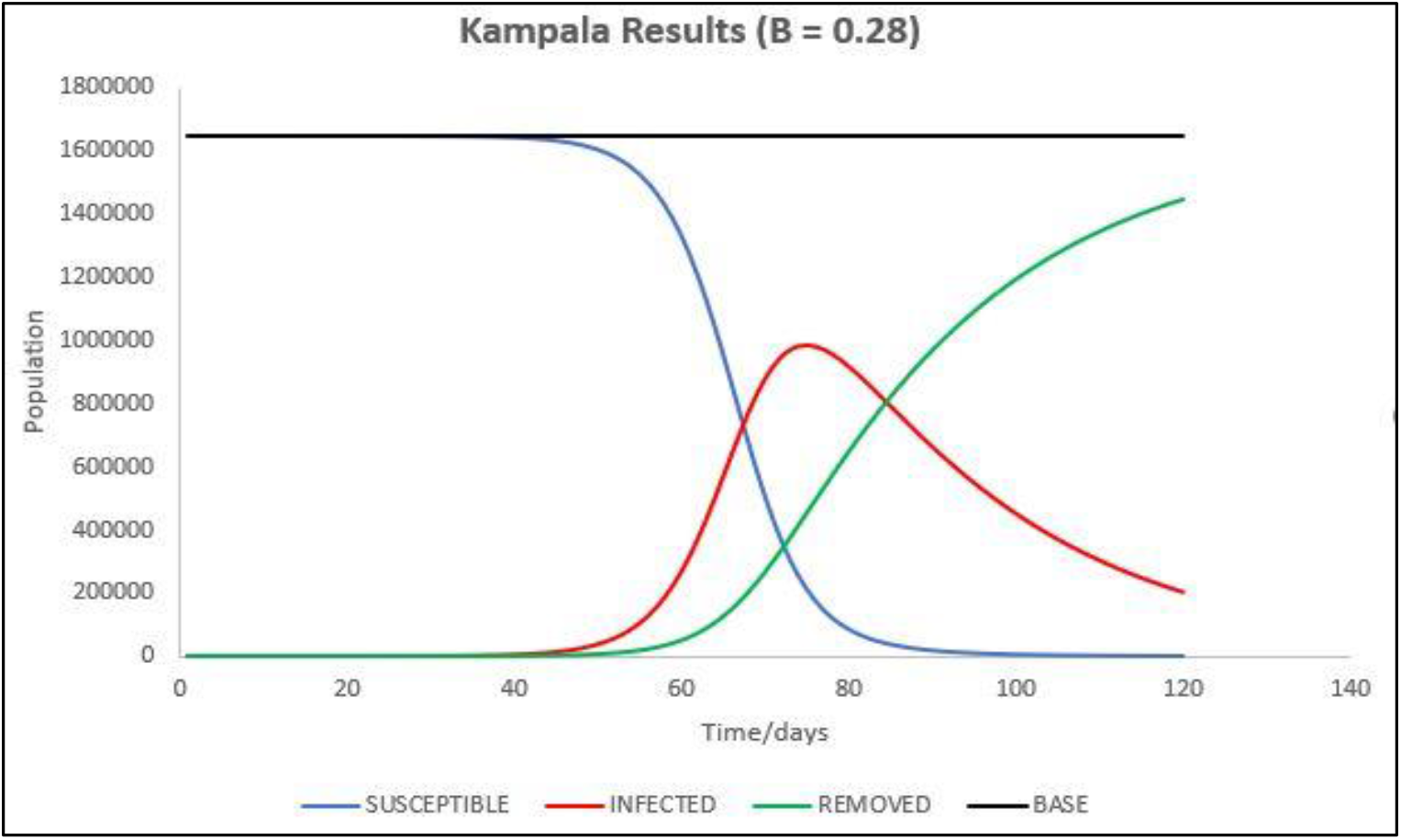
Results for Kampala assuming infection rate keeps at 0.28

**Figure 8:**
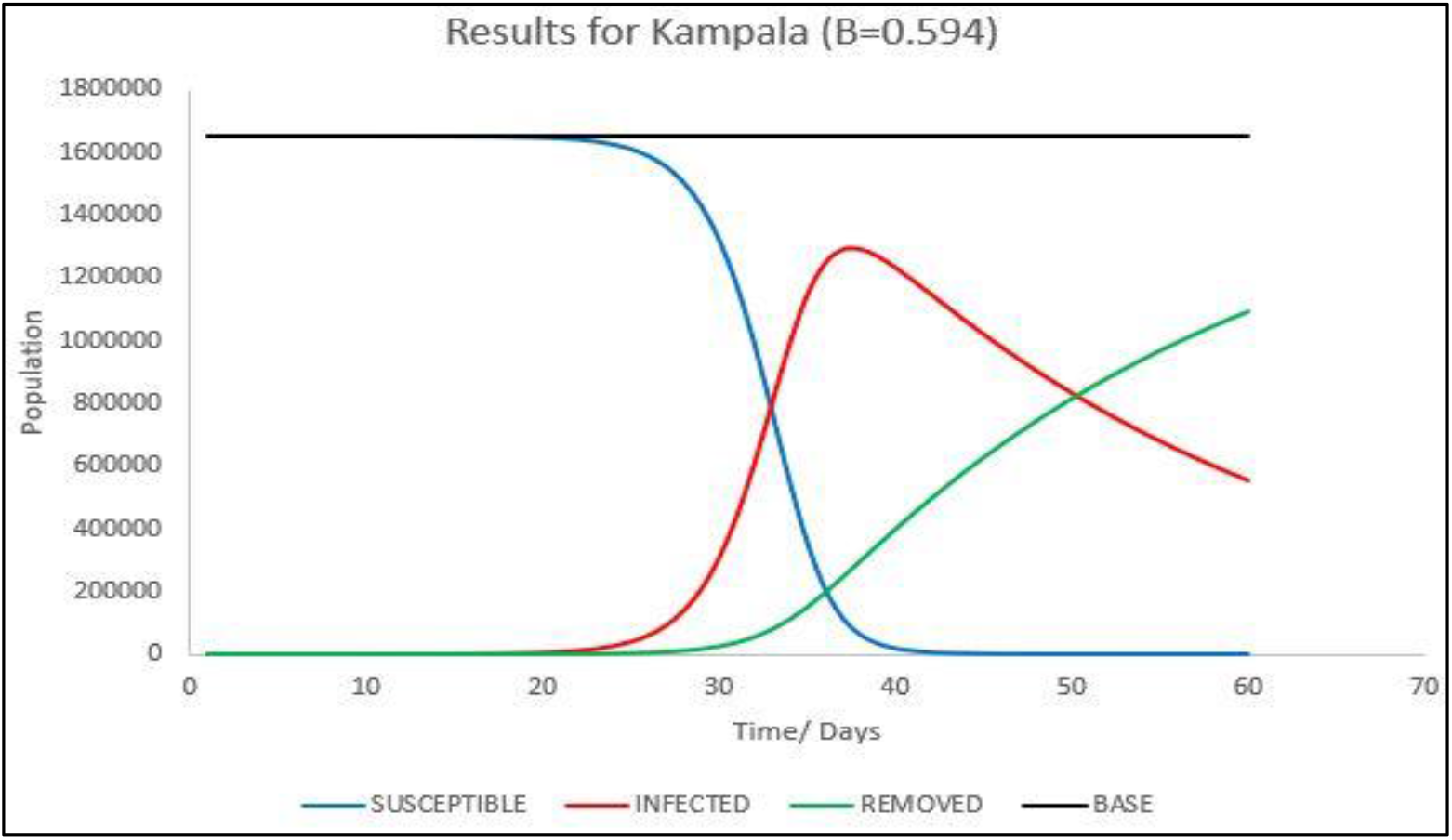
Excel Results for excel model if infection rate is 0.594

#### 3.3.4 DEATH PREDICTIONS

The ODE SIRmathematical excel model generated the results in figure1, considering the fact the SIR model doesn’t provide for provisions of a fatalities so every one in the population will be infected at some point, based on the number of days the vaccine stays in the place. We have remodeled the SIR to account for the number of deaths form infected individuals on a gjiven day.,a percentage of people is expected to die when they contract the disease and this is different for people in different age brackets as shown in figure. To match and predict individuals we need to split the number of infections to people in a given age bracket using probability theorems.

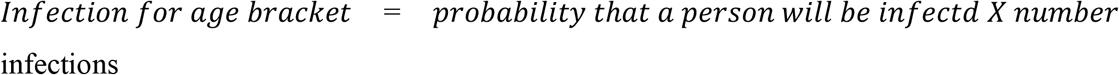

We have obtained the probability that a person of a particular age bracket will be infected as a fraction of number of people in that age bracket, sample demonstration is made based on the number of people per age bracket leaving in Kampala metropolitan area shown in the table below. For this test model we shall only predict the number of deaths in only one region (Kampala metropolitan area that includes people in Kampala and Wakiso districts, as per the data obtained from Uganda national Bureau of Statistics. Statistics were established on how the population in the region is distributed based on the age brackets of 2-20, 20 – 64, 64 and above as shown in the table below. The general death rate of all infected individuals around the globe is estimated to be at 3.4%. However, data generated from Chinese center for disease control estimates people of different age brackets to have different death rates as shown in the graph below. In our own predictions we match the death rates to the people living in Kampala metropolitan area. Assumption: the probability picking an individual in one of the age brackets is equal to the probability of picking someone who is infected and belongs to that age bracket. This would determine the possible number of people in a particular age bracket that will get an infection. Since the age brackets estimated by the CDC doesn’t match with the age brackets given by the Uganda National Bureau of Standards, we have harmonized the age brackets to just three and obtained the mean of the death rate of the range of people in that age bracket.

The first age bracket has people from the range of 2 to 20 years (young) there death rate is estimated at 0.2%, 20 to 64 years (adult) death rate is got from the mean of age brackets 20 to 29, 30 t0 39, 40 t0 49 and 50 to 64, in our model the four age brackets will be merged in one bracket 20 to 64. With a mean death rate 0.525% and all the other age brackets are catered for by the 65 plus (old) bracket with a mean death rate 8%, calculated the number of infections for three age brackets, and used the probailities of an infection choosing an individual given that he is from that age bracket.That probability multiplied by the number of total infectious individuals will generate the infections per particular age bracket.The average mortality rates we earlier on calculated for the four age brackets, will be multiplied multipled with the number of infectious individuals in that age bracked we are able to obtain the number of deaths for a given day.This applies the bayes theorem and conditional probabilities to generate the number of the death. Now we were able to determine, the number of deaths based on the consistence of the mortality per a particular age bracket. From the mortality rate ratios by the number of people infected in a population the first death is expected to occur on the, after the first death it means that the SIR model would have changed to account for the provision of the individual who died. So, the number of people infected would have decreased by one person. The trend in the infectious individuals would change by a certain function.

**Table 1:** shows the probability that a person in a particular age bracket will be infected, source Uganda national bureau of standards.

| AGE GROUP | Young | adult | elderly |
| --- | --- | --- | --- |
| POPULATION IN KMP | 4295035 | 784944 | 16913 |
| PERCENTAGE OF AGE<br>BRACKET | 0.84267 | 0.1540044 | 0.003318 |
| DEATH RATES | 0.2 | 0.525 | 8.8 |

**Figure 9:**
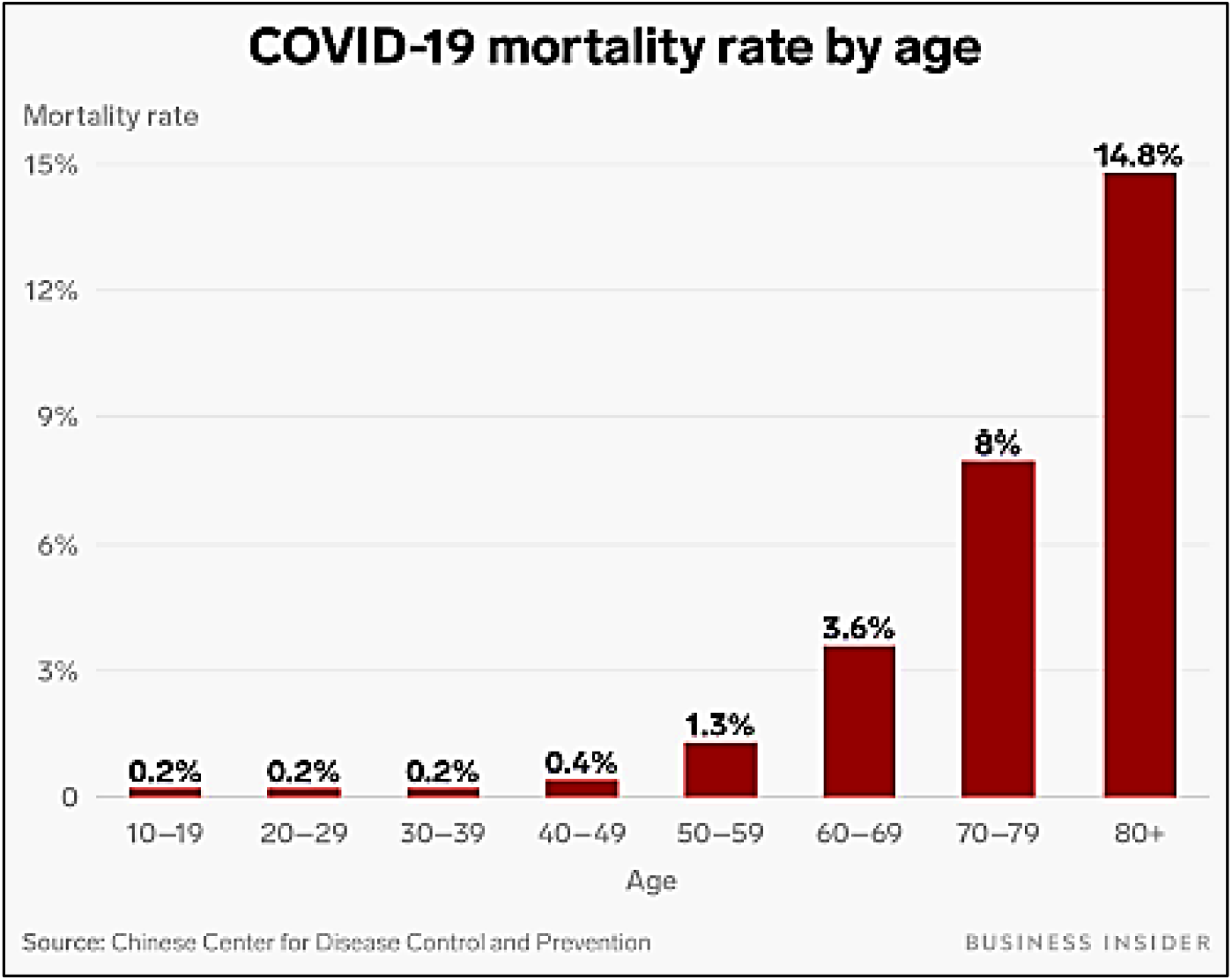
Death rates based on people of different age brackets from the Chinese center for disease control

#### 3.3.5 EXCEL WORK SHEETS

We have hence obtained worksheets for people who are like to be infected per a particular age brackets, who are directly extracted from the total number of people infected.

**Figure 10:**
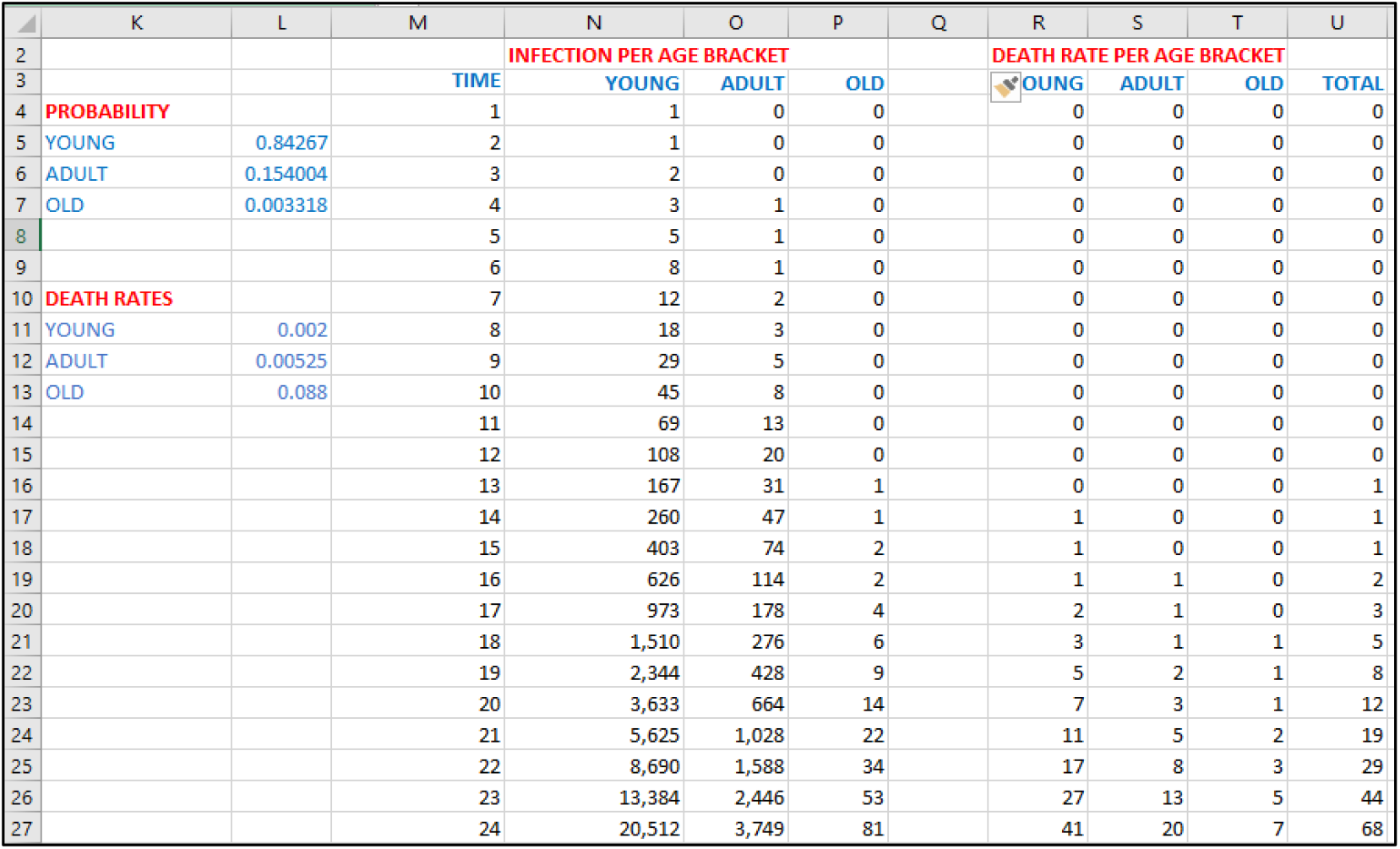
Excel worksheet that shows prediction of deaths from different age bracket

## RESULTS

From the excel worksheet above we can obtain cumulative graphs for total expected number of deaths in district like Kampala.

**Figure 11:**
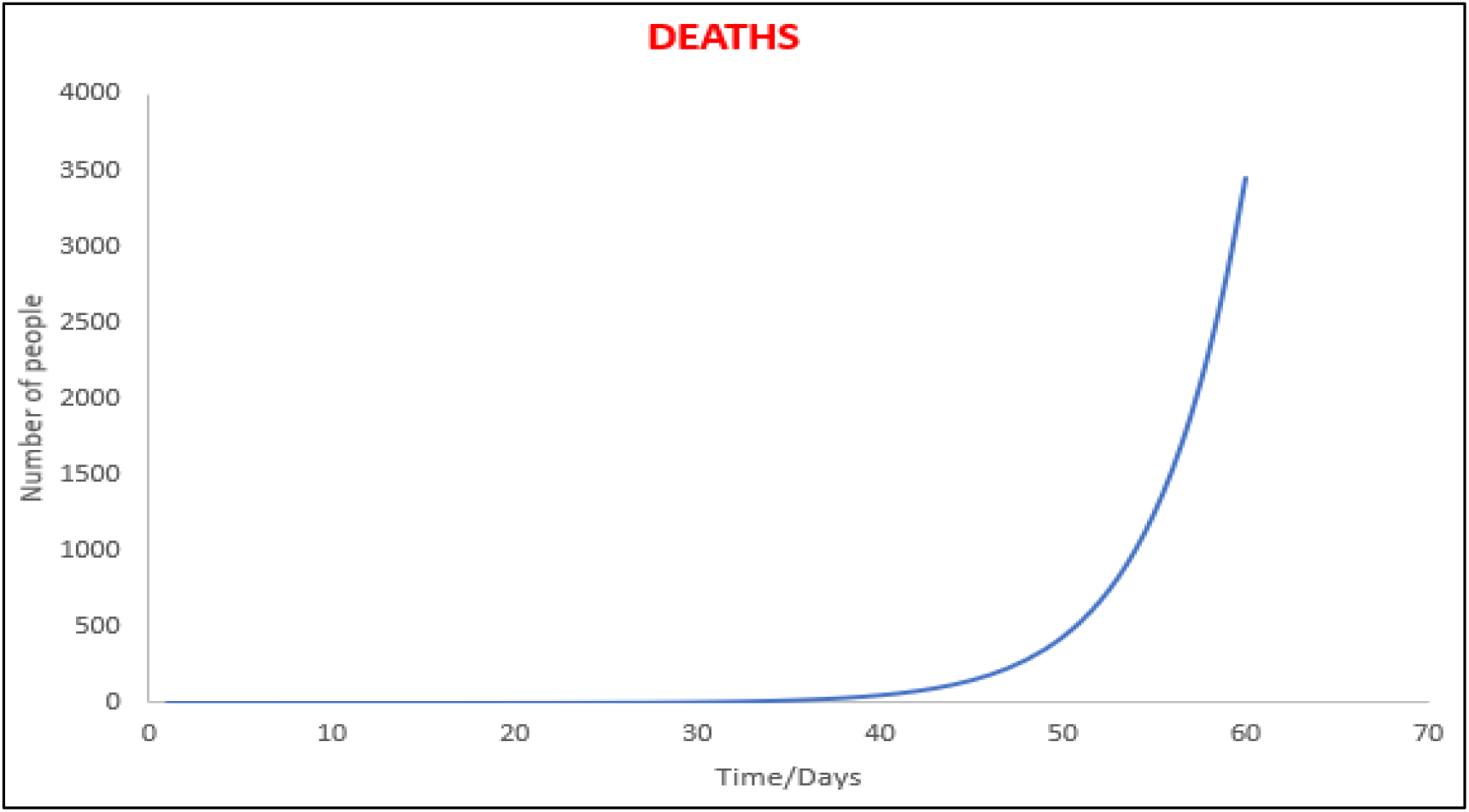
Graph showing predictive number of cumulative deaths in Kampala at B=0.28

## 4.0 DISCUSSION

For the excel and desktop application results simulation the rate of infection was varied at 0.594 a condition of infection for less government action and 0.28 for moderate government action. Results obtained in graphical format.

At B = 0.594

For this simulation a normal sustainable public transport do innated city of Kampala was assumed with very less government action to tackle the disease, for the first twenty days cases are very low and there is less rise mainly because of the incubation period, the sharp rise starts after the twentieth day and the epidemiological peak is reached at around the fourth day, by this time close to a million people may have been exposed or contracted the infection and it’s until the peak is reached, the cases begin to fall as more people begin to recover or die from the disease. There are no significant recoveries until the fourth day as more and more people start to recover.

At B = 0.28

This is assumed to be the case for uganda by this date were public transport is banned, the rate of infection is relatively lower than previous assumed rate, until the fourth day significant number of cases are not recorded and the cases sharply rise until the 70^th^ day were the epidemiological peak can be obtained for the curve to flatten, then cases begin to fall sharply as more people recover or die from the pandemic.

Deaths at B = 0.28

The simulations in excel showed that there were no deaths until day fourteen when the first death is recorder, the cases rise sharply to 500 by the 50th day and continue to rise until the peak is obtained, by 60^th^ day close to 3000 people are likely to have died from the disease.

## 5.0 CONCLUSION

By no means we cannot claim the accuracy of the results, due to varying dynamics of the disease the infection rates can change or vary at any time based on the actions taken by the government and populations. Both models sampled above produced relatively similar results with a few variations which may be based on numerical method estimations in excel model, the application above can be used to estimate rates for any given area with a few inputs and following the excel model procedure you can generate same results. The difference between the two models is ease of use, the application can easily be used by anyone, even with less computational skills unlike the excel model which may require prior expert knowledge with Microsoft excel, results predicted in both models may be accurate will smaller populations like for one city but can be in accurate when a big sample space is taken due to variance in dynamics. We hope these models can be employed in different areas and estimate how best to combat the deadly disease and allocate resources based on scientific and statistical facts and we further advise the use of both models at the same time, to get comparative results.

## Data Availability

there is one link to access of the code

